# Effects of wearing face masks under moderate physical effort

**DOI:** 10.1101/2021.03.18.21253539

**Authors:** Eberhart Martin, Orthaber Stefan, Kerbl Reinhold

## Abstract

Due to the COVID-19 pandemic, face masks have become part of daily life and work. Only few studies have investigated potential negative effects of this measure. It was aim of this study to evaluate whether face masks may affect oygen saturation and heart reate under moderate physical effort. Ten healthy volunteers were asked to climb up four floors (96 steps) with and without a face mask. Oxygen saturation and heart rate were recorded before and after this procedure.

Physical activity lead to a slight decrease of oxygen saturation of 3.0% (± 3.1) without and 3.7% (± 3.8) with face masks (p= 0.28). Heart rate increased by 31.0 (±18.7) and 33.3 (± 15.9) beats per minute (p= 0.39). Desaturation was more pronounced in individuals > 40 years, however was uncritical in all cases.

This study shows that in healthy adults the use of face masks does not significantly affect oxygen supply nor cause additional cardiac load.

## Introduction

Due to the COVID–19 pandemic, face masks or medical masks are recently recommended by international institutions (1,2). In several countries, the wearing of face masks is regulated by law for certain sectors of public life (3). Especially medical staff members and other health professionals are obliged to routinely wear face masks during work hours. Thus far, only few studies have investigated potential negative effects of this measure, although complaints of staff members about headache, fatigue, respiratory and other problems are frequent.

It was aim of this study to evaluate whether face masks may affect oygen saturation and heart reate under moderate physical effort.

## Patients and Methods

After approval by the local ethics committee (EK 33-108 ex 20/21), ten healthy volunteers (6 female, 4 male) were recruited to participate in this comparative study. Five subjects were younger than 30 years (mean 27±0.7), five older than 45 years (55.2±5.4). All subjects were in good health, non-smokers and used to wear masks during their daily work hours.

The participants were asked to climb up four floors (96 steps) in the same speed they would choose under routine conditions, once with (FM) and once without a face mask (NOFM). The order of the two runs was randomized and an interval of 5 minutes was scheduled in between. For FM runs, standard surgical face masks were used (CE norm DIN EN 14683). Oxygen saturation and heart rate were recorded immediately before and 10 seconds after physical activity by use of a pulse oximetric finger sensor (PULOX PO-200). Furthermore, the time spent for climbing the four floors was recorded.

Statistical analyses were done using Excel© (Microsoft Office, Excel 2013). For numerical data the t test was used. Statistical significance was set at p < 0.05.

## Results

Table 1 shows mean values of oxygen saturation and heart rate before and after physical activity for FM and NOFM runs, respectively. The mean time spent for climbing up the 4 floors was 65 (± 5) seconds for FM runs and 65 (± 6) for NOFM runs. Physical activity lead to a slight decrease of oxygen saturation of 3.7% (± 3.8) for FM and 3.0% (± 3.1) for NOFM runs (p= 0.28). Heart rate increased by 33.3 (± 18.7) and 31.0 (± 15.9) beats per minute, respectively (p= 0.39).

**Table 1:**
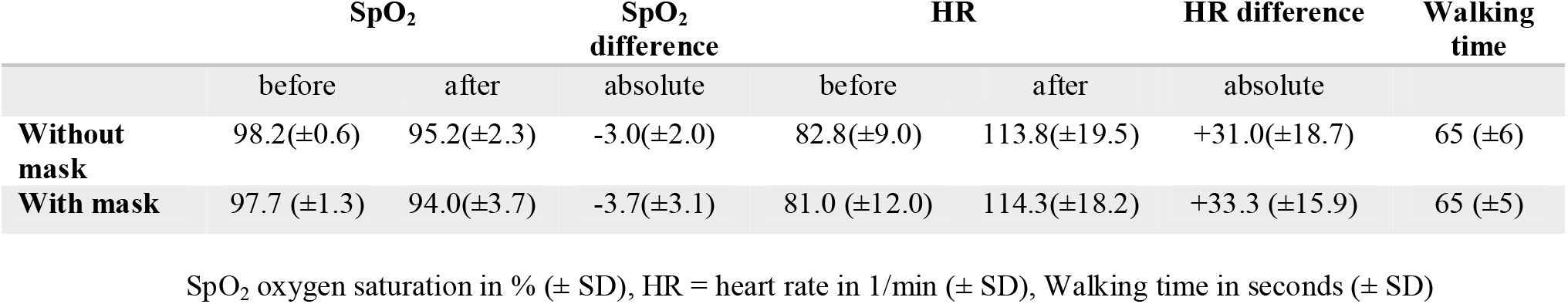
Mean values before and after moderate physical effort, n = 10.

For FM condition, desaturations were slightly more pronounced in individuals > 45 years than in those < 30 years (4.2±3.4 vs. 3.2±3.0, p=0.32, n.s.). In contrast, the increase of HR was less prominent in individuals > 45 years for FM condition (27.6 ± 19.5 vs. 39.0 ± 10.4; p = 0.141, n.s.).

## Discussion

Our study demonstrates that in healthy adults face masks do not negatively affect oxygen supply and cardiac function under conditions of moderate physical activity. These findings are in line with previous studies that report no significant drop in oxygen saturation and, in particular, no potentially harmless alterations (4–8). Our recordings demonstrate that moderate physical activity (climbing up 96 steps) leads to a moderate decrease of oxygen saturation, similarly with and without face mask.

Beder et al. found that a postoperative SpO_2_ decrease was more pronounced in surgeons over the age of 35 when compared to younger colleagues (7). In our study, the older age group also showed a higher drop of SpO_2_ when using a mask, the difference was however not statistically significant.

Wearing a face mask did not significantly influence the HR in our study, neither in rest nor after moderate physical activity. Roberg et al. report a slightly higher HR for subjects walking on a treadmill and wearing face masks (5). In contrast, Butz did not note any compensatory increase of HR through face mask use (6). Dattel et al. similarly found no influence upon HR (8). In addition, these authors also recorded end-tidal CO_2_ values (etCO_2_), which proved to remain in the normal range in flight instructors wearing face masks for 90 minutes (8).

Wearing a mask did not change walking speed while climbing up four floors. This corresponds to the results of Person et al. who demonstrated that walking distance remains unchanged in a six-minute walking test when wearing a mask (9).

The number of studies dealing with medical masks is limited. Most of these studies were performed in rest or under light physical activity such as walking. The HR increase of 31/min without a mask in our study shows that climbing four floors represents a moderate short-term exercise. The effects of full physical stress were investigated by Fikenzer et al. during spiroergometry in healthy persons (10). The results of this study showed that ventilation and cardiopulmonary training capacity were reduced by surgical masks, especially by FFP2/N95 masks.

The sample size of our study was relatively low (n=10). The results were however quite consistent in all volunteers and lead to the conclusion that surgical masks do not negatively affect oxygen supply and cardiac function during moderate physical activity.

Our results suggest that complaints reported as a consequence of wearing a face mask are not due to hypoxemia or cardiac load. In contrast, surgical masks appear to be safe for healthy adults even under conditions of moderate physical activity.

## Data Availability

Raw data were generated at LKH Hochsteiermark. Derived data supporting the findings of this study are available from the corresponding author on request.

## Declarations

### Institutional Review Board Statement

The study was conducted according to the guidelines of the Declaration of Helsinki, and approved by the Ethics Committee of Graz (EK 33-108 ex 20/21 from 4.12.2020)

### Ethical standards

All procedures followed were in accordance with the ethical standards of the responsible committee on human experimentation (institutional and national) and with the Helsinki Declaration of 1975, as revised in 2008. Approval was given by local ethics committee (EK 33-108 ex 20/21),

### Informed Consent Statement

Informed consent was obtained from all patients for being included in the study.

## Authors’ contributions

Martin Eberhart: methodology, investigation, writing - original draft, project administration Stefan Orthaber: resources, data curation, visualization Reinhold Kerbl: conceptualization, validation, writing - review & editing, supervision

## References

[1] European Centre for Disease Prevention and Control. Using Face Masks in the Community - Reducing COVID-19 Transmission from Potentially Asymptomatic or Pre-Symptomatic People through the Use of Face Masks.; 2020. Accessed November 20, 2020. https://www.ecdc.europa.eu/en/publications-data/using-face-masks-community-reducing-covid-19-transmission

[2] Robert Koch-Institut. Mund-Nasen-Bedeckung im öffentlichen Raum als weitere Komponente zur Reduktion der Übertragungen von COVID-19. Strategie-Ergänzung zu empfohlenen Infektionsschutzmaßnahmen und Zielen (3. Update). Epid Bull. 2020;19:3–5. doi:10.25646/6731

[3] Verordnung des Bundesministers für Soziales, Gesundheit, Pflege und Konsumentenschutz, mit der die COVID-19-Maßnahmenverordnung geändert wird (3. COVID-19-MV-Novelle) vom 22.10.20 https://www.ris.bka.gv.at/Dokumente/BgblAuth/BGBLA_2020_II_455/BGBLA_2020_II_455.pdfsig-abegrufenam25.10.20

[4] Samannan R, Holt G, Calderon-Candelario R, Mirsaeidi M, Campos M. Effect of face masks on gas exchange in healthy persons and patients with COPD. Ann Am Thorac Soc. Published online 2020. doi:10.1513/AnnalsATS.202007-812RL

[5] Roberge RJ, Kim J-H, Benson SM. Absence of consequential changes in physiological, thermal and subjective responses from wearing a surgical mask. Respir Physiol Neurobiol. 2012;181(1):29–35.

[6] Butz U. Rückatmung von Kohlendioxid bei Verwendung von Operationsmasken als hygienischer Mundschutz an medizinischem Fachpersonal. Tum.de. Accessed November 23, 2020. https://mediatum.ub.tum.de/doc/602557/602557.pdf

[7] Beder A, Büyükkoçak U, Sabuncuoğlu H, Keskil ZA, Keskil S. Preliminary report on surgical mask induced deoxygenation during major surgery. Neurocirugia (Astur). 2008;19(2):121–126.

[8] Dattel AR, O’Toole NM, Lopez G, Byrnes KP. Face mask effects of CO2, heart rate, respiration rate, and oxygen saturation on instructor pilots. Collegiate Aviation Review International. 2020;38(2):1. Retrieved from http://ojs.library.okstate.edu/osu/index.php/CARI/article/view/8038/7412

[9] Person E, Lemercier C, Royer A, Reychler G. Effect of a surgical mask on six minute walking distance. Rev Mal Respir. 2018;35(3):264–268.

[10] Fikenzer S, Uhe T, Lavall D, et al. Effects of surgical and FFP2/N95 face masks on cardiopulmonary exercise capacity. Clin Res Cardiol. doi:10.1007/s00392-020-01704-y

